# Cross-sectional and Longitudinal Associations Between Circadian Alignment and Cardiovascular-Kidney-Metabolic Syndrome in US Adults

**DOI:** 10.1101/2025.03.25.25323796

**Authors:** Bolin Li, Xinping Kang, Yushan Liu, Li Ma, Qing Zhang, Lele Cheng, Kai Lan, Yongxin Yang, Jialing He, Miaomiao Cao, Zhen Zhang, Rong Jie

**Affiliations:** Department of Cardiology, Affiliated Hospital of SouthwestJiaotong University, The Third People’s Hospital of Chengdu, Chengdu, 610014, Sichuan, China; Dermatology, Xi’an Jiaotong University First Affiliated Hospital, Xi’an, Shaanxi 710004, China; Xi’an Jiaotong University, Xi’an, Shaanxi 710004, China; Intensive Care Unit, Bazhong Central Hospital, Bazhong, 636000, Sichuan, China; Department of Medical Imaging, the First Affiliated Hospital of Xi’an Jiaotong University, Xi’an, Shaanxi 710004, China; The Affiliated Hospital of Shaanxi University of Chinese Medicine, Xianyang, Shaanxi 712046, China

**Author notes:** Denotes joint first author (Bolin Li, Xinping Kang). Denotes joint corresponding author; Correspondence: Rong Jie,.

**Keywords:** Circadian, Cardiovascular-kidney-metabolic syndrome, mortality, NHANES

## Abstract

**Background:** Cardiovascular-kidney-metabolic (CKM) syndrome is a progressive, multi-systemic condition defined by recent consensus. Circadian rhythm is a critical regulator of metabolic health, but the relationship between circadian alignment and CKM remains unclear.

**Objective:** To explore the association between circadian alignment and CKM risk as well as mortality outcomes.

**Methods:** NHANES data from 2011-2014 were used. Circadian alignment was evaluated using phasor analysis of light-activity synchronization, including phasor magnitude and acrophase, categorized into quintiles. CKM stages (0-4) were used to reflect disease progression, comparing advanced stages (3 or 4) with non-advanced stages (0, 1, or 2). Outcomes included advanced CKM stages, cardiovascular disease (CVD), diabetes mellitus (DM), chronic kidney disease (CKD), all-cause mortality, cardiovascular mortality, and premature mortality. Advanced CKM stages, CVD, DM, and CKD were analyzed cross-sectionally, while mortality outcomes were assessed longitudinally.

**Results:** A total of 7,246 participants, representing approximately 148 million US adults (median follow-up: 81 months), were included. Participants in the lowest quintile of phasor magnitude, indicating the greatest circadian misalignment, had significantly higher risks of advanced CKM stages (aOR 2.25, 95% CI: 1.59 – 3.19), all-cause mortality (aHR 1.75, 95% CI: 1.35–2.26), cardiovascular mortality (SHR 1.89, 95% CI: 1.22–2.93) compared to those in the highest quintile. However, premature mortality was not significantly associated. A linear inverse relationship was observed between phasor magnitude and both advanced CKM stages and mortality outcomes, with similar patterns for CVD, CKD, and DM. No significant associations were found between phasor acrophase and adverse outcomes.

**Conclusions:** Greater circadian misalignment, indicated by lower phasor magnitude, is strongly associated with an increased risk of advanced CKM and other adverse outcomes. Improving circadian alignment may offer a promising strategy for mitigating the burden of cardiovascular and metabolic diseases.

## Introduction

Cardiovascular-kidney-metabolic (CKM^1^) syndrome stresses the pathophysiological interplay between metabolic risk factors, chronic kidney disease (CKD), and cardiovascular disease (CVD)(1), affecting 90% of U.S. adults. To profile syndrome drivers and countermeasures, the AHA stratified CKM staging (0-4) designates Stage 3-4 as advanced disease(2). CKM syndrome results in a great economic burden and has emerged as the primary cause of mortality(3).

Circadian rhythm operates on a roughly 24-hour cycle, coordinating the timing of physiological processes to keep synchrony with environmental cues, primarily the light-dark cycle. These oscillations regulate sleep architecture, hormone release(4), metabolism(5,6), eating habits(7), or digestion. Its disruption from lifestyle, shift work, or genetic factors triggers systemic desynchronization, elevating risks of metabolic-cardiovascular-renal disorders(8,9).

Diabetes mellitus (DM) and metabolic syndrome patients exhibit disrupted circadian rhythms, which correlate with elevated risks of hypertension (10) and cardiovascular complications(8). Also, researchers demonstrated the impacts of circadian rhythm disruption on renal physiology, including glomerular filtration efficiency, tubular reabsorption, abnormal sodium handling, renin-angiotensin - aldosterone system dynamics, and compromising systemic blood pressure regulation(9), indicating bidirectional circadian-CKM interactions. Population-level quantification of intricate relationship between circadian rhythm disruptions and CKM may provide strategies for modifiable prevention of CKM syndrome.

Here, we collected data from the NHANES study cohort, using phasor analysis to examine the associations of circadian alignment and CKM risk and mortality outcomes, aiming to identify the prospective targets for CKM management.

## Method

### Data source and study population

Data in the present investigation were retrieved from the National Health and Nutrition Examination Survey (NHANES^2^), which is a national, serial, one-time probability sampling design program to track health and nutrition patterns across age groups. Approval for the NHANES protocol was granted by the National Center for Health Statistics Ethics Review Board. The ethics approval was not required for this study because it was a secondary analysis based on a publicly available database and did not include identified personal information. The present research adhered to the Strengthening the Reporting of Observational Studies in Epidemiology (STROBE) for reporting observational studies in epidemiology, specifically for cross-sectional studies.

NHANES data from the two-year cycles of 2011-2012, 2013-2014 are screened in this study. Of all 19,931 participants in the 2011-2014 NHANES survey, we excluded those aged < 20 years and lacking information of CKM syndrome determination, survival outcome, and light and/or activity data. Consequently, a total of 7493 participants were included.

### Phasor analysis

Light exposure and physical activity patterns were objectively monitored were measured by physical activity monitoring devices in NHANES survey(11). Phasor analysis evaluates the 24-hour light intensity and motility fluctuations by two variables including phasor magnitude and acrophase(12). Light exposure has a nonlinear impact on the human circadian system (13). To address circadian photoreception nonlinearity, light intensity data were adjusted via logistic curves with a 10,000 lux biological ceiling.

Circadian stimulation (CS^3^) metrics from Daysimeter recordings underwent logistic transformation to map threshold-to-saturation responses(12). Behavioral entrainment was assessed through circular cross-correlations, revealing timing-related relationships. Normalization involved dividing by total samples, adjusting by standard deviations, ensuring correlation coefficients range from −1 to 1(14). This analysis provides a robust framework for understanding circadian stimulation’s impact on behavioral responses, utilizing statistical methods to extract meaningful correlations from complex datasets. The phasor magnitude indicates the extent of alignment between an individual’s light-dark cycle and his/her activity cycle, while the phasor acrophase reflects the temporal relationship between the two cycles.

### Definition of CKM and other conditions

CKM syndrome stages were categorized according to modified criteria based on Aggarwal et al(2), adapted for NHANES data fields: Stage 0: BMI 18.5-24.9 kg/m^2^ and waist circumference below 88 cm (women) or 102 cm (men), with no higher stage criteria met. Stage 1: BMI ≥25 kg/m^2^, WC ≥88 cm (women) or ≥102 cm (men), or prediabetes (fasting glucose 100-124 mg/dL, HbA1c 5.7-6.4%, or antidiabetic therapy). Stage 2: Presence of metabolic risk factors (triglycerides ≥ 135 mg/dL, hypertension, diabetes, or metabolic syndrome), or moderate/high-risk CKD(15). Stage 3: Very high-risk CKD or elevated 10-year CVD risk (16). Stage 4: History of CVD. CKM syndrome was further classified as advanced (stages 3-4) or non-advanced (stages 0-2)(2) CVD was defined as history of coronary heart disease, angina, heart attack, heart failure, and stroke. CKD risk is classified based on estimated glomerular filtration rate(GFR^4^) classification and albumin levels, with moderate risk serving as the threshold for albuminuria as suggested(1,15), with moderate risk serving as the threshold for defining CKD as a binary variable. DM was determined as HbA1c≥6.5%, fasting glucose>7.0mmol/L, self-reported diagnosis of diabetes, or usage of diabetes medications.

### Outcomes

Outcomes included advanced CKM stages, CVD, DM, CKD, all-cause mortality, cardiovascular mortality, and premature mortality. Advanced CKM stages, CVD, DM, and CKD were analyzed cross-sectionally, while mortality outcomes were assessed longitudinally.

### Covariates

Potential confounders, including age as a continuous variable, gender (female or male), racial and ethnic background (Hispanic; non-Hispanic Black, non-Hispanic White, and other [multiracial and any race other than Black or White]), smoking behavior (current, former, <100 cigarettes in life or never smoker), and alcohol use (nondrinker, < 1/week, 1/week to 1/day, 1/day above), education (less than high school, high school graduate, some college, college graduate and above or others), marital status(marital, unmarried or unknown), household income(<20000, 20000-44900, 45000-74900, >75000 and unknown), and Body Mass Index(BMI)(<18.5, 18.5-25, 25-30, ≥30). All the covariates are self-reported, mutually exclusive categories. Demographic characteristics and lifestyle habits were obtained by personal interview, and BMI was derived from physical measurements at the Mobile Examination Center.

### Statistic analysis

Analytical approaches incorporated CDC-recommended NHANES sampling weights (WTINT2YR) to account for complex survey design effects, including cluster sampling and non-response bias. All statistical procedures adhered to CDC analytic guidelines for complex survey data. Quintiles grouped phasor magnitude and angle to characterize the research population. Continuous measures reported as weighted mean ± standard deviation (normal) or medians with interquartile range (skewed), categorical variables as adjusted proportions. Group comparisons utilized Kruskal Wallis, and Rao-Scott χ^2^ tests. The cross-sectional analysis of the relationship between phasor magnitude and advanced CKM stages utilizes binary logistic weighted regression. Longitudinal analyses addressed cause-specific mortality rate, based on Cox proportional hazards regression models. Model 1 was adjusted for age and sex using logistic regression, with additional adjustment for advanced CKM stages in the Cox regression. Model 2 further controlled for other confounding variables such as race and ethnicity, education level, marital status, and household income. Model 3 further accounted for smoking, alcohol consumption, and BMI. Additionally, the Fine and Gray method was applied to estimate the competing risks in cardiovascular mortality. And follow-up period had a median of 81 months (interquartile range [IQR] 70-95). To assess the potential linear relationship between phasor magnitude and adverse outcomes, restricted cubic splines were applied, with adjustment for the covariates in Model 3.

All our statistical analyses used R4.4.1 (R Foundation for Statistical Computing, Vienna, Austria) statistical software, except for competing risk regression analysis was conducted using STATA 17.0 based on the Fine and Gray model.

## Results

### Baseline characteristics of participants

Descriptive data by phasor magnitude quintile are shown in **Table 1**. A total of 7,246 participants represented 148,082,293 U.S. adults (**Supplementary Figure 1**). Among them, 48% were male, 68% non-Hispanic white, with a weighted median age of 50 years. The median phasor magnitude and acrophase were 0.36 (0.28-0.43) and 0.70 (−0.11-1.57), respectively. The study revealed that 31% had advanced CKM stages, 12% (n=3,231,405) had CKD, 17% (n=4,316,430) had CVD, and 15% (n=4,068,029) had DM.

**Table 1.** Baseline Demographic and Clinical Characteristics of the Study Population.

|  | Phasor magnitude<sup>1</sup> |  |  |  |  | *P* value |
| --- | --- | --- | --- | --- | --- | --- |
|  | Q1 | Q2 | Q3 | Q4 | Q5 |  |
| Phasor magnitude,median(IQR) | 0.20 (0.16, 0.23) | 0.29 (0.27, 0.31) | 0.35 (0.33, 0.36) | 0.41 (0.39, 0.42) | 0.48 (0.46, 0.51) | <0.001 |
| Phasor Acrophase,median(IQR) | 0.91 (-0.69, 2.34) | 0.89 (0.09, 1.79) | 0.82 (-0.01, 1.57) | 0.62 (-0.07, 1.44) | 0.46 (-0.14, 1.10) | <0.001 |
| Age(%) |  |  |  |  |  | <0.001 |
| 20-39 | 9,816,924 (37%) | 10,096,082 (35%) | 9,013,348 (31%) | 8,673,912 (28%) | 7,930,076 (24%) |  |
| 40-59 | 10,234,358 (39%) | 10,961,091 (38%) | 10,966,586 (38%) | 12,945,061 (41%) | 13,104,961 (40%) |  |
| 60+ | 6,284,067 (24%) | 7,520,784 (26%) | 8,907,132 (31%) | 9,731,169 (31%) | 11,896,744 (36%) |  |
| gender(%) |  |  |  |  |  | 0.011 |
| female | 12,276,843 (47%) | 14,207,489 (50%) | 16,174,014 (56%) | 17,280,928 (55%) | 17,108,556 (52%) |  |
| male | 14,058,505 (53%) | 14,370,467 (50%) | 12,713,052 (44%) | 14,069,213 (45%) | 15,823,226 (48%) |  |
| Race and ethnicity(%) |  |  |  |  |  | <0.001 |
| Hispanic | 1,403,312 (5.3%) | 1,683,691 (5.9%) | 1,795,080 (6.2%) | 2,015,566 (6.4%) | 1,862,293 (5.7%) |  |
| Non-Hispanic Black | 4,633,728 (18%) | 3,668,122 (13%) | 3,263,076 (11%) | 2,669,875 (8.5%) | 1,489,527 (4.5%) |  |
| Non-Hispanic White | 16,255,486 (62%) | 18,621,752 (65%) | 19,341,389 (67%) | 22,047,092 (70%) | 24,573,591 (75%) |  |
| Other | 4,042,823 (15%) | 4,604,392 (16%) | 4,487,521 (16%) | 4,617,609 (15%) | 5,006,370 (15%) |  |
| education level(%) |  |  |  |  |  | <0.001 |
| college graduate or above | 8,758,461 (33%) | 10,244,292 (36%) | 9,749,853 (34%) | 9,642,357 (31%) | 8,258,101 (25%) |  |
| high school graduate | 5,085,669 (19%) | 5,082,898 (18%) | 5,959,426 (21%) | 6,300,516 (20%) | 8,371,905 (25%) |  |
| Less than high school | 3,463,955 (13%) | 3,320,454 (12%) | 4,365,792 (15%) | 5,434,091 (17%) | 6,520,862 (20%) |  |
| some college | 8,987,676 (34%) | 9,925,815 (35%) | 8,811,994 (31%) | 9,964,724 (32%) | 9,780,913 (30%) |  |
| missing | 39,588 | 4,496 | 0 | 8,454 | 0 |  |
| marriage(%) |  |  |  |  |  | <0.001 |
| Marital | 12,353,211 (47%) | 15,173,493 (53%) | 16,238,690 (56%) | 19,465,343 (62%) | 21,392,410 (65%) |  |
| not married | 13,982,138 (53%) | 13,404,463 (47%) | 12,648,376 (44%) | 11,884,798 (38%) | 11,532,385 (35%) |  |
| missing | 0 | 0 | 0 | 0 | 6,986 |  |
| household income(%) |  |  |  |  |  | 0.032 |
| <20,000 | 4,419,523 (17%) | 4,105,840 (15%) | 4,199,173 (15%) | 4,352,958 (14%) | 4,503,324 (14%) |  |
| 20,000–44,900 | 8,237,377 (32%) | 6,925,288 (25%) | 7,761,724 (28%) | 8,376,772 (28%) | 10,186,478 (32%) |  |
| 45,000–74,900 | 4,098,034 (16%) | 5,950,178 (21%) | 5,626,140 (20%) | 6,601,103 (22%) | 6,604,534 (21%) |  |
| 75,000+ | 8,917,827 (35%) | 10,733,356 (39%) | 10,366,385 (37%) | 10,965,539 (36%) | 10,847,880 (34%) |  |
| missing | 662,587 | 863,293 | 933,644 | 1,053,769 | 789,566 |  |
| BMI(%)<sup>2</sup> |  |  |  |  |  | 0.4 |
| elevated | 19,249,109 (74%) | 20,222,947 (71%) | 20,915,749 (73%) | 23,649,332 (76%) | 24,043,734 (73%) |  |
| normal | 6,900,390 (26%) | 8,176,237 (29%) | 7,857,435 (27%) | 7,654,959 (24%) | 8,888,047 (27%) |  |
| missing | 185,850 | 178,771 | 113,881 | 45,851 | 0 |  |
| Smoke(%) |  |  |  |  |  | <0.001 |
| current | 6,123,432 (23%) | 5,161,282 (18%) | 4,695,329 (16%) | 5,719,500 (18%) | 6,329,003 (19%) |  |
| former | 4,543,657 (17%) | 6,724,608 (24%) | 7,680,548 (27%) | 8,605,511 (27%) | 9,770,327 (30%) |  |
| no | 15,659,549 (59%) | 16,682,649 (58%) | 16,506,229 (57%) | 17,017,758 (54%) | 16,818,045 (51%) |  |
| missing | 8,710 | 9,416 | 4,959 | 7,373 | 14,407 |  |
| Drink(%) |  |  |  |  |  | <0.001 |
| 1/day above | 624,593 (2.4%) | 1,128,046 (3.9%) | 841,568 (2.9%) | 1,777,901 (5.7%) | 2,871,126 (8.7%) |  |
| 1/week to <1/day | 7,238,767 (27%) | 8,801,689 (31%) | 9,184,558 (32%) | 9,414,505 (30%) | 9,883,451 (30%) |  |
| <1/week | 9,522,003 (36%) | 10,324,606 (36%) | 9,562,381 (33%) | 10,371,964 (33%) | 10,477,470 (32%) |  |
| no | 8,949,986 (34%) | 8,323,616 (29%) | 9,298,558 (32%) | 9,785,772 (31%) | 9,699,734 (29%) |  |
| CKM_stage |  |  |  |  |  | 0.15 |
| Stage-0 | 2,718,809 (10%) | 3,310,526 (12%) | 2,808,886 (9.7%) | 2,711,976 (8.7%) | 2,888,008 (8.8%) |  |
| Stage-1 | 5,702,962 (22%) | 5,637,875 (20%) | 5,238,536 (18%) | 6,141,368 (20%) | 6,010,415 (18%) |  |
| Stage-2 | 12,770,224 (48%) | 14,857,412 (52%) | 15,902,197 (55%) | 17,527,027 (56%) | 19,051,928 (58%) |  |
| Stage-3 | 1,911,948 (7.3%) | 1,911,131 (6.7%) | 1,795,716 (6.2%) | 2,108,797 (6.7%) | 1,730,290 (5.3%) |  |
| Stage-4 | 3,231,405 (12%) | 2,861,011 (10%) | 3,141,730 (11%) | 2,860,974 (9.1%) | 3,251,141 (9.9%) |  |
| CVD<sup>3</sup> | 3,231,405 (12%) | 2,861,011 (10%) | 3,141,730 (11%) | 2,860,974 (9.1%) | 3,251,141 (9.9%) | 0.3 |
| CKD<sup>4</sup> | 4,316,430 (17%) | 4,269,661 (15%) | 4,155,896 (14%) | 4,682,719 (15%) | 4,260,640 (13%) | 0.4 |
| DM | 4,068,029 (15%) | 4,301,013 (15%) | 4,037,728 (14%) | 4,533,223 (14%) | 4,199,979 (13%) | 0.4 |
| Follow time (month) | 81 (70, 95) | 80 (70, 95) | 82 (69, 95) | 85 (71, 96) | 84 (71, 97) | 0.12 |

1. All percentages and medians (IQRs) are weighted using sample weight
2. normal body mass index (BMI) (<23 kg/m<sup>2</sup> for individuals with Asian ethnicity and <25 kg/m<sup>2</sup> for other racial and ethnic groups)
elevated BMI (≥23 kg/m<sup>2</sup> for individuals with Asian race and >25 kg/m<sup>2</sup> for all other race and ethnic groups)
3. CVD was identified based on self-reported established cardiovascular disease (coronary heart disease, angina, heart attack, heart failure, and stroke)
4. CKD was identified based on Kidney Disease Improving Global Outcomes (KDIGO) criteria, with CKD risk as moderate-to-very-high risk.

### Association between circadian alignment and the risk of advanced CKM stages

Firstly, we evaluated the relationship of phasor variables (magnitude and acrophase) and advanced CKM risk. Three logistic regression models were built to identified the association between phasor magnitude and advanced CKM risk (**Figure 1A**). Model 1 shows stepwise increased trends for advanced CKM risks with reduced phasor magnitude. Specifically, participants in Q1 group (the lowest magnitude) are 252% (odds ratio [OR] 2.52, 95% CI 1.86-3.14) more likely to have advanced CKM compared with the highest magnitude group (Q5 group). Although further adjusted for potential confounders slightly attenuated the association in model 2 and 3, the first quartile (Q1) group always demonstrated highest risk of advanced CKM, with an OR of 2.25 (95% CI: 1.59-3.19) in the fully adjusted model 3. **Figure 1B-D** also revealed that shorter phasor magnitude exhibited a higher risk for each CVD (OR 1.66, 95% CI 1.66-2.23), CKD (OR 1.62,95% CI 1.14-2.30) and DM risk(OR 1.47, 95% CI 1.11-1.93) in the fully adjusted model 3, individually.

**Figure 1.**
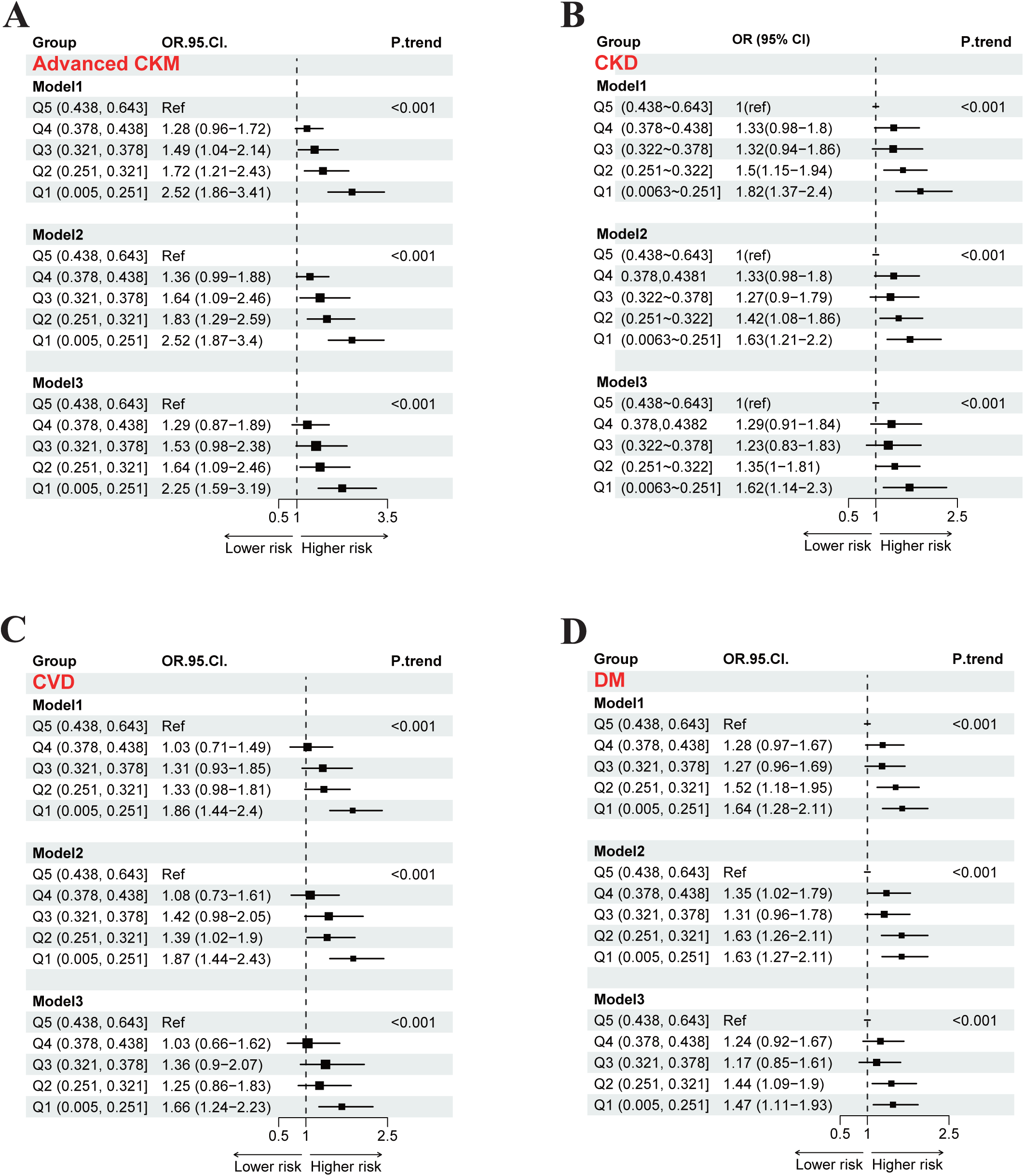
Association between phasor magnitude and CKM risk (A), CKD risk (B), CVD risk (C) and DM risk(D). All plots are performed using multivariate logistic regression based on Model 1 (age and sex), Model 2 (additional race and ethnicity, education level, marital status, and household income), Model 3(additional smoking, alcohol consumption, and BMI). ORs and 95% CI are presented comparing with Q1 (referrence group). BMI: body mass index.

The restricted cubic spline (RCS) analysis revealed a linear negative correlation between continuous phasor magnitude and the risk of advanced CKM, as illustrated in **Figure 2A** based on Model3. Similar correlations were observed in CVD, CKD, and DM, as shown in **Figures 2D, 2G**, and **2J**. Furthermore, subgroup analyses were conducted to identify the correlations between continuous phasor magnitude and the risks of advanced CKM and each separate CKM conditions, stratified by sex (female, male) and age (grouped by 20-39, 40-59, 60 and beyond). The curves showed similar linear negative correlations across different subgroups, as shown in **Figure 2**. Interestingly, male and older individuals with short phasor magnitude were more likely to have advanced CKM and each separate CKM condition, compared with their female and younger counterparts.

**Figure 2.**
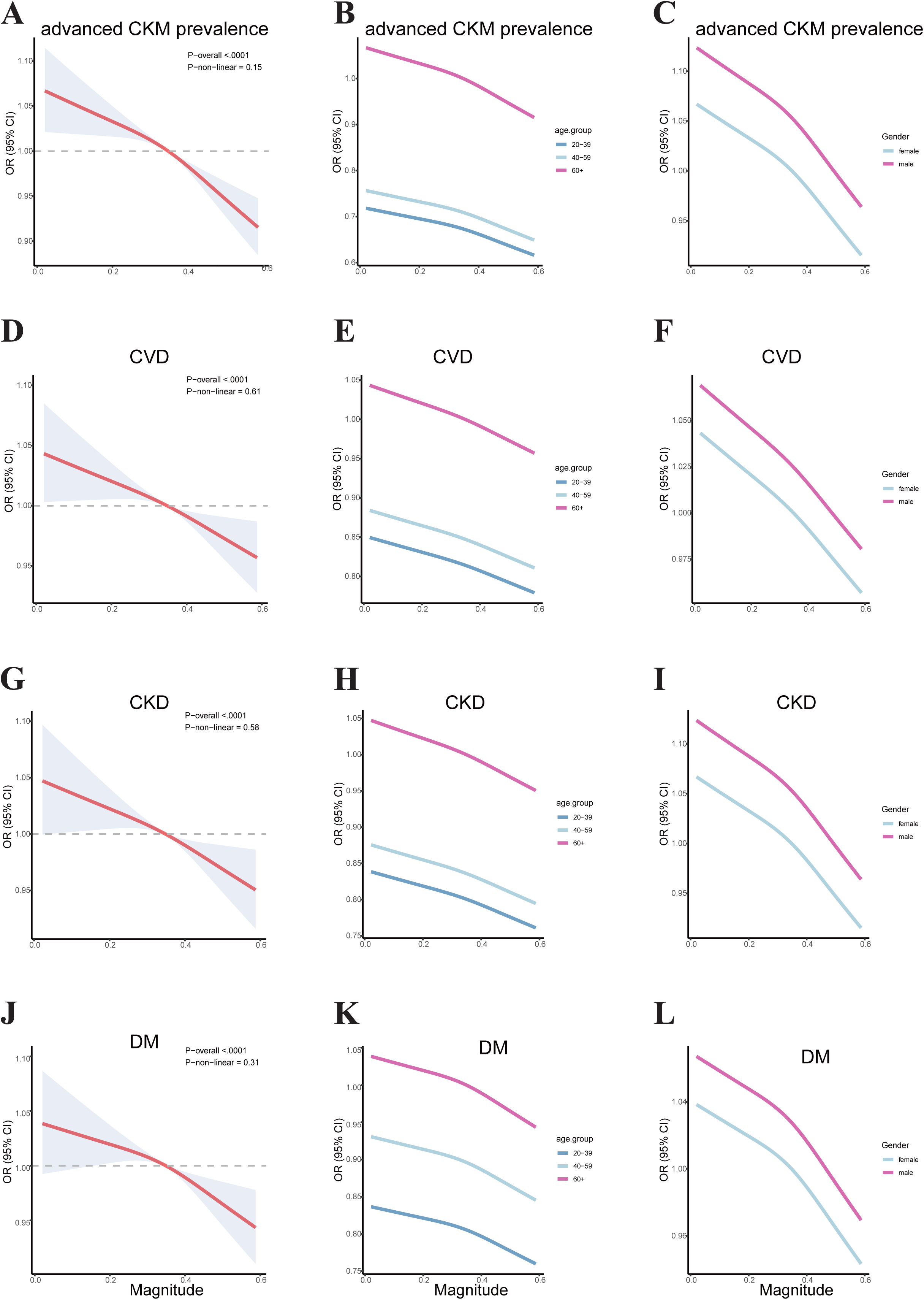
Restricted cubic spline curves for association between phasor magnitude and CKM* risk (A-C), CVD* risk (D-F), CKD* risk (G-I) and DM* risk(J-L). All plots are performed using multivariate logistic regression based on Model 3. The hazard ratio is represented by a red line and the 95% confidence interval is represented by the shaded area.The right two plots of each row illustrate subgroup analyses adjusted for sex and age.CKM: cardiovasculer-kidney-metabolic syndrome; CKD: chronic-kidney disease CVD: cardiovascular disease; DM: diabetes mellitus.

There was no statistical significance between acrophase quintiles and the advanced CKM risk, as well as CVD, CKD, and DM risks in the adjusted logistic regression models (**supplementary Figure 2**). However, RCS curves indicated U-shaped associations between acrophase and advanced CKM, CVD, in which small and large acrophase were related to higher CKM and CVD risks compared with median acrophase. Moreover, acrophase was negatively correlated with CKD risk, and positively correlated with DM risk (**supplementary Figure 3**). Similar to phasor magnitude, male and older individuals exhibited higher risks of advanced CKM and each separate CKM conditions, compared with their female and younger counterparts in the subgroup analyses (**supplementary Figure 3**).

### Association between circadian alignment and mortality outcomes

We investigated the relationship between phasor variables and mortality outcomes, including all-cause, cardiovascular, and premature mortality. The cumulative hazard function was used to assess risks for adverse outcomes across magnitude quartile groups, as shown in **Figure 3**. During a median follow-up of 81 months (interquartile range 70-95), lower phasor magnitudes were associated with a poorer prognosis, particularly in the first magnitude quartile (Q1). The heatmap reveals that mortality increases with the number and overlap of conditions (CVD, CKD, DM) and decreases with higher phasor magnitudes (**Supplementary Figure 4**). Additionally, the proportion of adverse outcomes appeared to rise with the number of conditions (**Supplementary Figure 4**).

**Figure 3.**
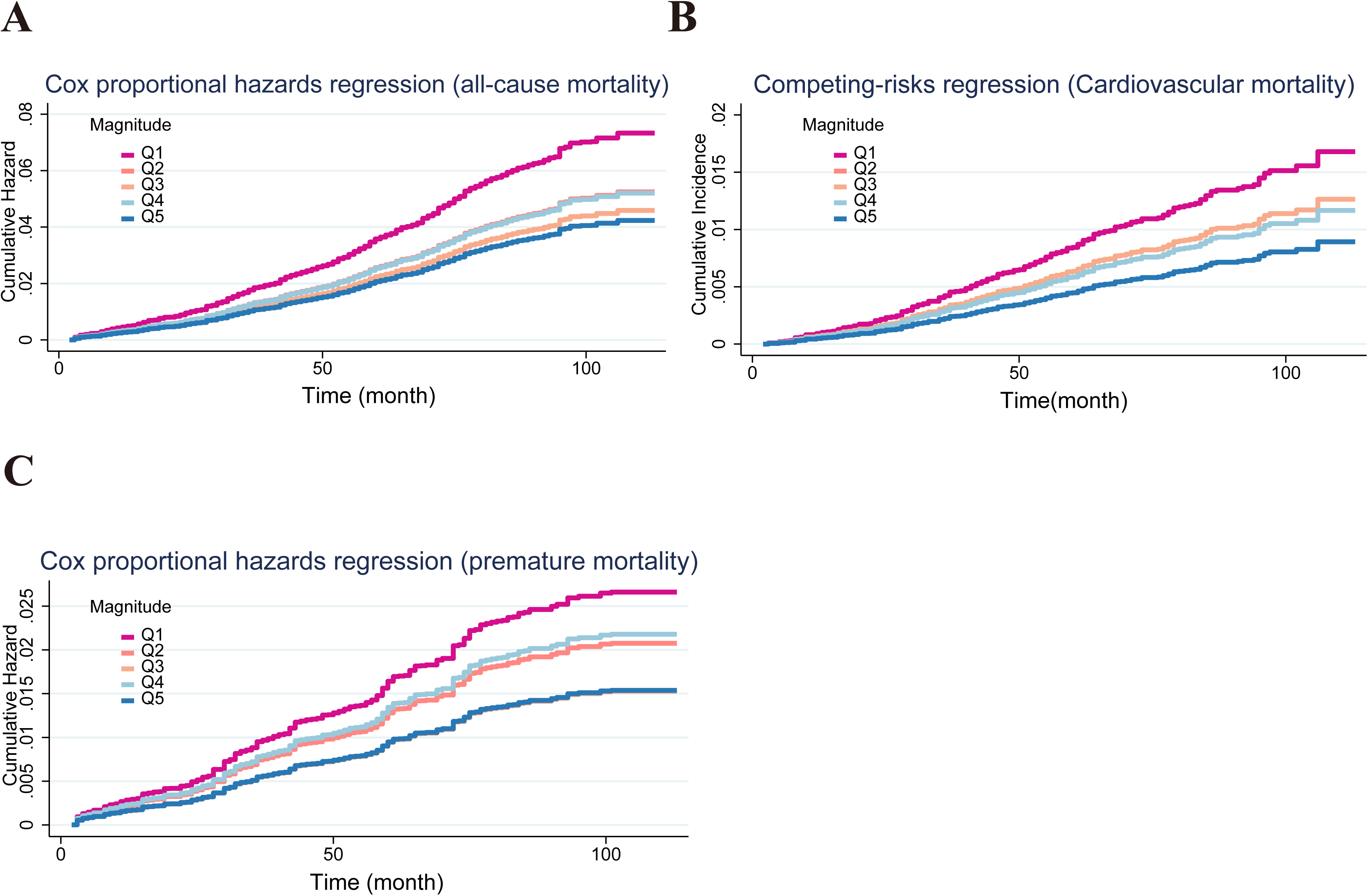
Cumulative hazard function curves competing risks regression analysis under Fine and Gray models estimates for selected endpoints, cardiovascular mortality(B), cox (all-cause mortality(A), and premature mortality(C)) by phasor magnitude. All plots are performed using Cox proportional hazards regression based on Model 3.

In the Cox proportional hazards models, the Q1 group was independently associated with increased all-cause mortality (aHR 1.46, 95% CI 1.11 –1.94) and cardiovascular mortality (SHR 1.84, 95% CI 1.21–2.81) after adjusting for advanced CKM stages and other confounders in model 1. This relationship remained significant in models 2 and 3 (**Figures 4A-B**). Notably, in the fully adjusted Model 3, the Q1 group has a 75% and 89% higher risk of all-cause mortality and cardiovascular mortality, respectively, compared to the Q5 group (**Figure 4B**). No significant association was found between phasor magnitude and premature mortality (**Figure 4C**). RCS analyses in **Figure 5** confirmed a linear negative correlation between continuous phasor magnitude and mortality outcomes, including all-cause and cardiovascular mortality, as well as premature mortality. Stratification by sex, age, and advanced CKM status revealed consistent trends for all-cause and cardiovascular mortality, with higher risks observed in males, older individuals, and those with advanced CKM, compared to their counterparts.

**Figure 4.**
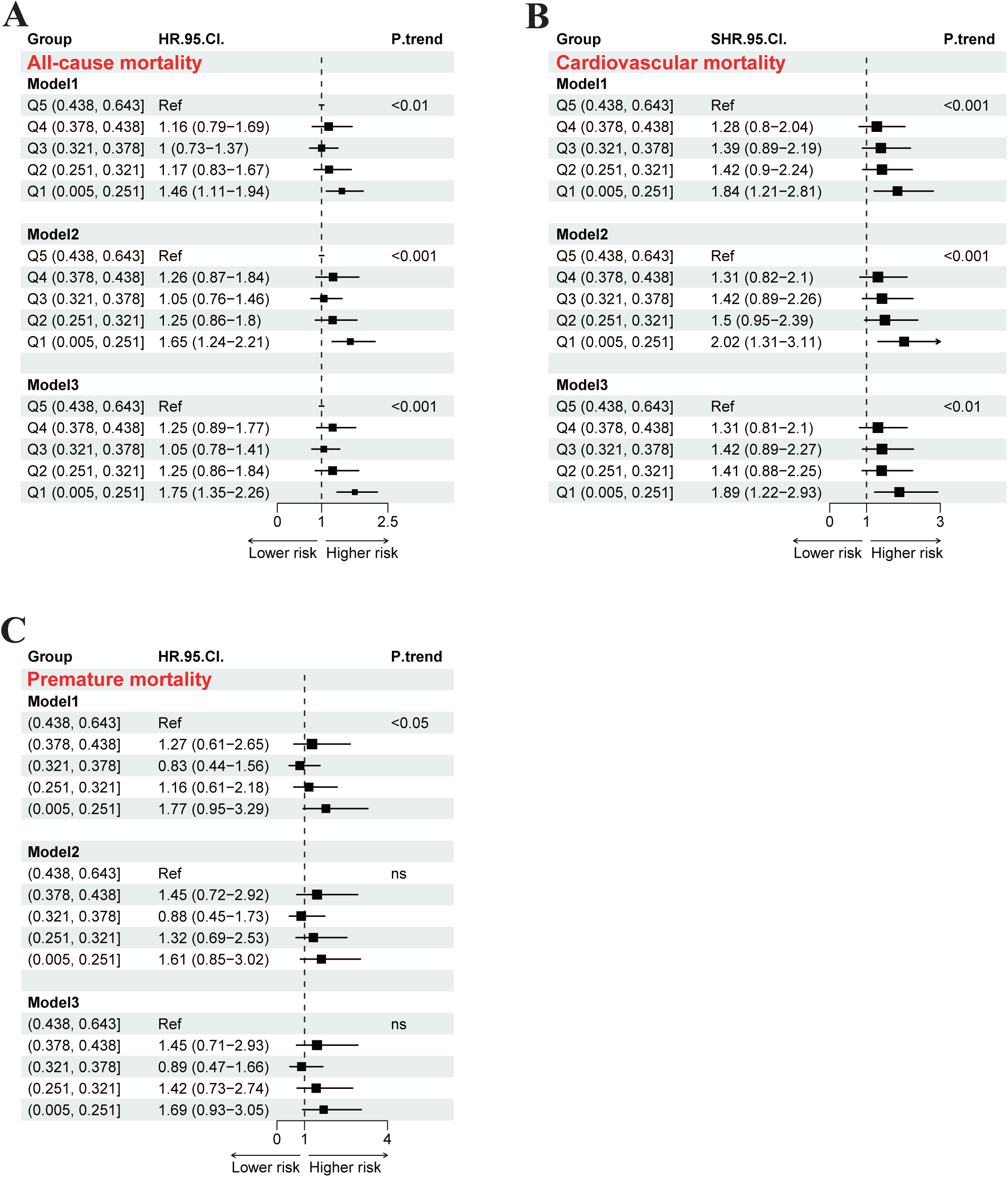
Association between phasor magnitude and all-cause mortality (A), cardiovascular mortality (B), premature mortality (C). Plots are performed using multivariate Cox proportional hazards regression - Model 1 (age, sex, and advanced CKM stages), Model 2 (additional race and ethnicity, education level, marital status, and household income), Model 3(additional smoking, alcohol consumption, and BMI). HRs and 95% CI are presented comparing with Q1 (referrence group).

**Figure 5.**
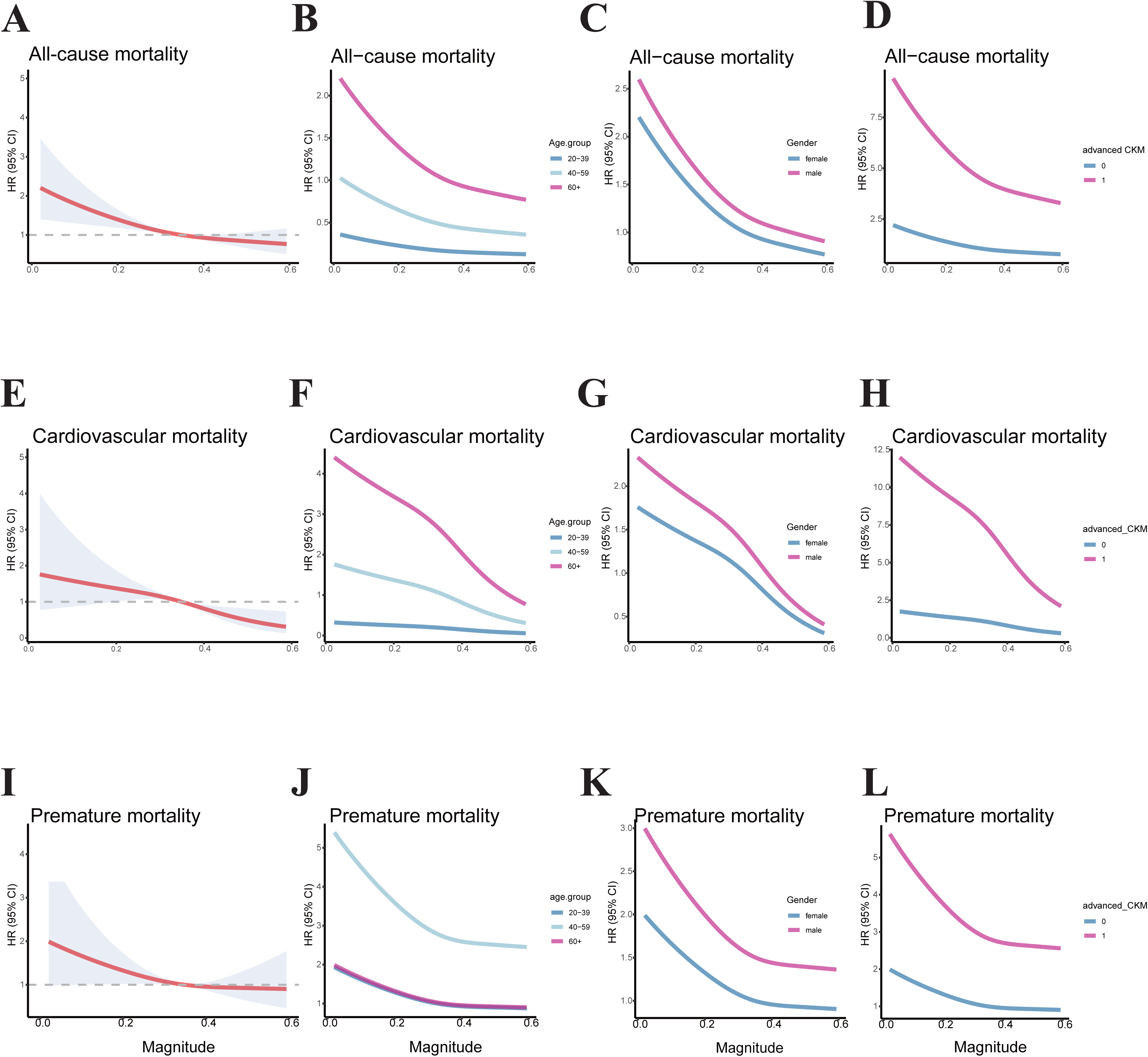
RCS curves for association between phasor magnitude and all-cause mortality (A-D), cardiovascular mortality (E-H), and premature mortality (I-L). All plots are performed using multivariate Cox proportional hazards regression based on Model 3. The hazard ratio is represented by a red line and the 95% confidence interval is represented by the shaded area. The right 3 plots of each row illustrate subgroup analyses stratified by sex, age and the number of ckm conditions.

However, no significant association was identified between phasor acorphase and adverse outcomes (**Supplementary Figure 5**), despite a linear correlation for all-cause, cardiovascular, and premature mortality (**Supplementary Figure 6**).

## Discussion

This cross-sectional and longitudinal study investigates the association between circadian alignment, CKM risk, and outcomes in U.S. adults using a nationally representative sample. The results indicate that phasor magnitude (the coupling extent of activity and light) is inversely related to the risk of advanced CKM stages, and adverse mortality outcomes, including all-cause and cardiovascular mortality. Additionally, males and older adults exhibited higher CKM incidence and all-cause/cardiovascular mortality compared to their female and younger counterparts. Participants with a greater number of coexisting conditions, including CVD, CKD and DM, also showed higher frequencies of all-cause and cardiovascular mortality. The most pronounced differences were observed among those with shorter phasor magnitudes.

In essence, circadian dysfunction and CKM are both significant public health issues(17,18)mediated by bidirectional interactions between endogenous physiology (cellular/behavioral processes) and exogenous environmental cues(19–21). Whereas existing research predominantly examines isolated circadian metrics (light-dark cycles or activity-rest patterns) or transient misalignments (e.g., chronotypes/social jet lag)(20), our study employs phasor analysis to quantify continuous light-activity cycle coupling, a validated biomarker of circadian dysregulation (11,14,22). Our observations align with Xiao et al.’s report that reduced phasor magnitude predicts incident diabetes mellitus, correlating with elevated HOMA-IR, OGTT, and fasting insulin levels (11). Growing evidence also showed that circadian dysfunction has an impact on CVD and CKD. Two large-scale population studies revealed insomnia associated with an increased risk for acute myocardial infarction(23) and incident heart failure(24). Nighttime artificial light exposure and late-night dietary intake exhibit quantifiable associations with heightened metabolic syndrome predisposition and elevated cardiovascular morbidity risk(25). A Mendelian randomization study including 465,814 UK Biobank participants demonstrated short sleep duration (<6 hours) was significantly related to a higher risk of CKD stages 3–5(26). Critically, diminished phasor magnitude predicts graded risks of CVD, CKD, and CKM severity, underscoring its translational utility for identifying high-risk populations.

Existing studies have examined inadequate sleep to elevated all-cause mortality (HR 1.23, 95% CI 1.13–1.34) and cardiovascular deaths (HR 1.39, 1.19–1.62) versus restorative sleep, with low physical activity exacerbating risks in a UK Biobank study (27). This association can be exacerbated by low physical activity. Further, in diabetic populations, healthy sleep patterns similarly reduce all-cause mortality(28), aligning with our finding that minimal phasor magnitude (light-activity misalignment) independently predicts higher all-cause/cardiovascular mortality. A Pooled analysis of 958,674 subjects across 16 cohorts confirms rotating/night shifts elevate all-cause/cardiovascular mortality versus daytime schedules(29). Furthermore, our analysis demonstrated a similar inverse association between phasor magnitude and mortality rates within the CKM cohort, a pattern consistently observed in CVD-related comorbidities and additional CKM-related health states, CKD. Wu et al. suggested that disorders including DM, coronary artery disease, and depression, partially mediate the causal relationship that short sleep duration was negatively correlated with lifespan(30). Our findings further identifies that circadian misalignment (shorter phasor magnitude) and CKM multimorbidity jointly amplify mortality risk.

Interestingly, male and older individuals were more likely to have CKM as well as present with higher risks of all-cause and cardiovascular mortality, especially for those with short magnitude. Indeed, sex and age-related disparities for circadian and CKM have been well-recognized. The “lateness” of chronotypes reaches a maximum around age 20, and then shift earlier with increasing age. In comparison with women, men show their “lateness” of chronotypes later and maintain most of their adulthood. Individuals aged over 60 years old mostly showed early chronotype(20,31). Men and people aged over 65 have higher prevalence of advanced CKM stages(2). Although the underlying mechanisms have not been elucidated, interaction of multiple factors, such as hormonal levels, 24-hour gene expression rhythms, and social factors et al., might involve in the different vulnerabilities to circadian dysfunction, CKM risk and outcomes between sex and age.

Regarding phasor acorphase, there was no statistical significance between acorphase and CKM risk and outcomes. Because the acrophase of light and activity peaks may be shifted forward or backward. Positive and negative acrophases may offset each other, thereby weakening their association with diseases and adverse outcomes.

Our study has some limitations. A key limitation of this observational design is the inability to establish causality between CKM-related risk factors and the coherence of the activity cycle and light cycle. Some studies also show cardiovascular disease can lead to circadian rhythm disorder, and vice versa. Second, we did not rule out all potential confounding factors that could influence the results, like seasons, genetic susceptibility, and underlying health conditions. Third, individuals with more severe diseases and hospitalized patients may have regular routines due to physician recommendations, leading to high magnitude; however, their disease severity and mortality rates are elevated. Fourth, we didn’t consider other daily activities such as physical exercise and meal timing, which are likely related to health status.

Reduced alignment between circadian rhythms and daily activity patterns independently predicts incident cardiometabolic-kidney multimorbidity (CKM) and adverse outcomes. Our findings, integrating novel phasor analysis with competing-risk models, position circadian optimization as a modifiable target for primary prevention— particularly in males and older adults, who face disproportionate CKM burdens.

Public health strategies should prioritize scalable circadian interventions (e.g., community-based light exposure guidelines, age-tailored physical activity programs) to curb the rising global burden of metabolic-kidney disorders. Mechanistic trials evaluating circadian-aligned therapies in high-risk populations are urgently needed.

## Data Availability

All data produced are available online at https://wwwn.cdc.gov/nchs/nhanes/default.aspx

## Acknowledgements

The authors would like to thank Dr. Lele Cheng and Dr. Qing Zhang for for their support in funding acquisition.

## Informed Consent Statement

All NHANES participants provided written informed consent prior to data collection.

## Data Availability Statement

The datasets analyzed during the current study are publicly available from the U.S. Centers for Disease Control and Prevention NHANES repository. Derived analytic code and processed data can be obtained from the corresponding author upon reasonable request.

## Authors’ contribution

Conception and design: Rong Jie, Bolin Li. Conducted research: Rong Jie, Miaomiao Cao. Formal analysis: Zhen Zhang, Kai Lan, Ma Li. Vusualisation: Yushan Liu, Yongxin Yang. Funding acquisition: Lele Cheng, Qing Zhang. Project administration: Zhen Zhang. Software: Rong Jie, Jialing He. Methodology: Bolin Li, Xinping Kang, Lele Cheng. Writing-original draft: Xinping Kang, Bolin Li. Review &editing: Miaomiao Cao, Bolin Li.

## Footnotes

1 CKM: Cardiovascular-kidney-metabolic syndrome

2 NHANES: National Health and Nutrition Examination Survey

3 CS: circadian stimulation

4 GFR: glomerular filtration rate

