## Supplementary figures and images for "Cross-sectional and Longitudinal Associations Between Circadian Alignment and Cardiovascular-Kidney-Metabolic Syndrome in US Adults"

### supplement_Page1.tif

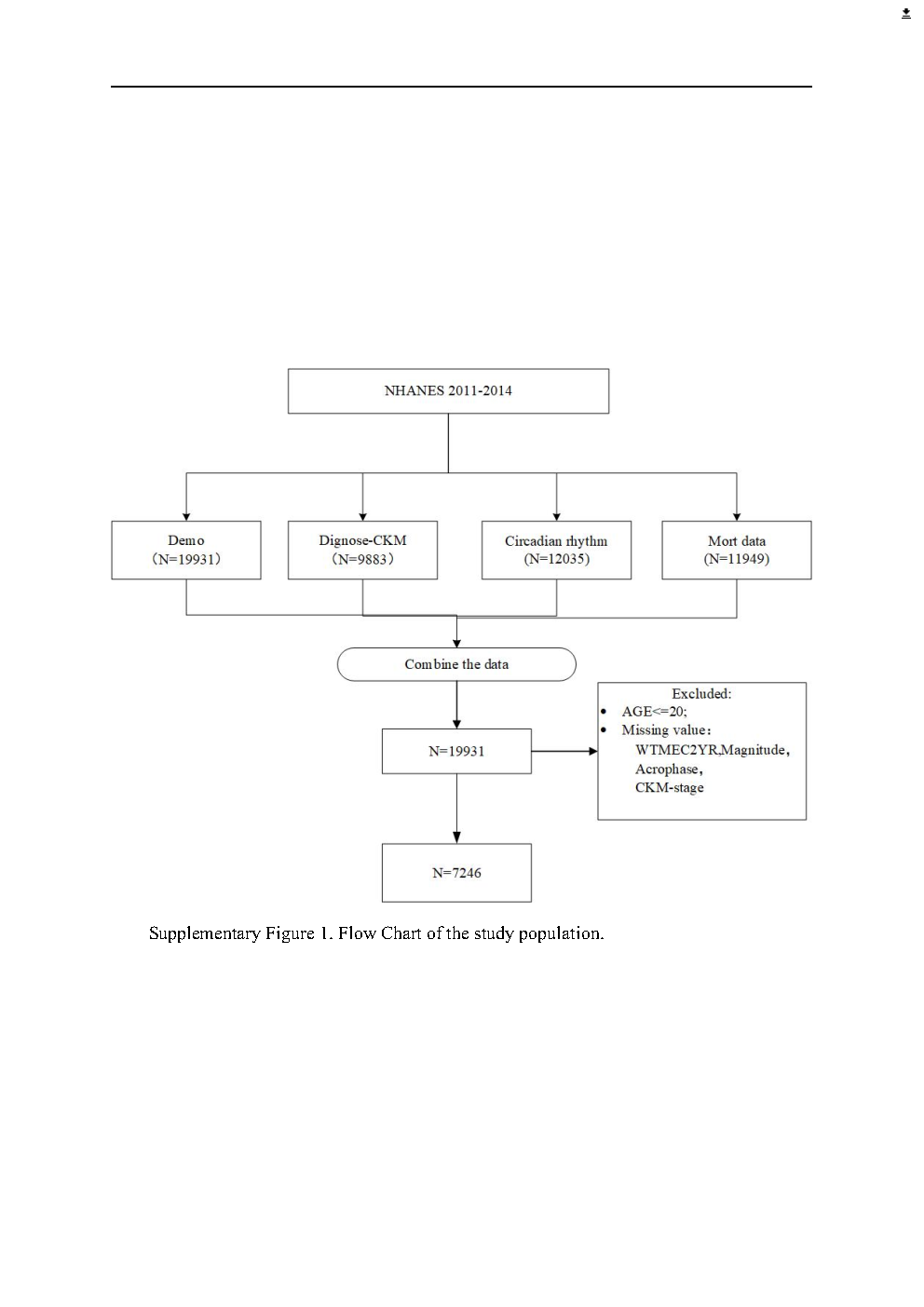

### supplement_Page2.tif

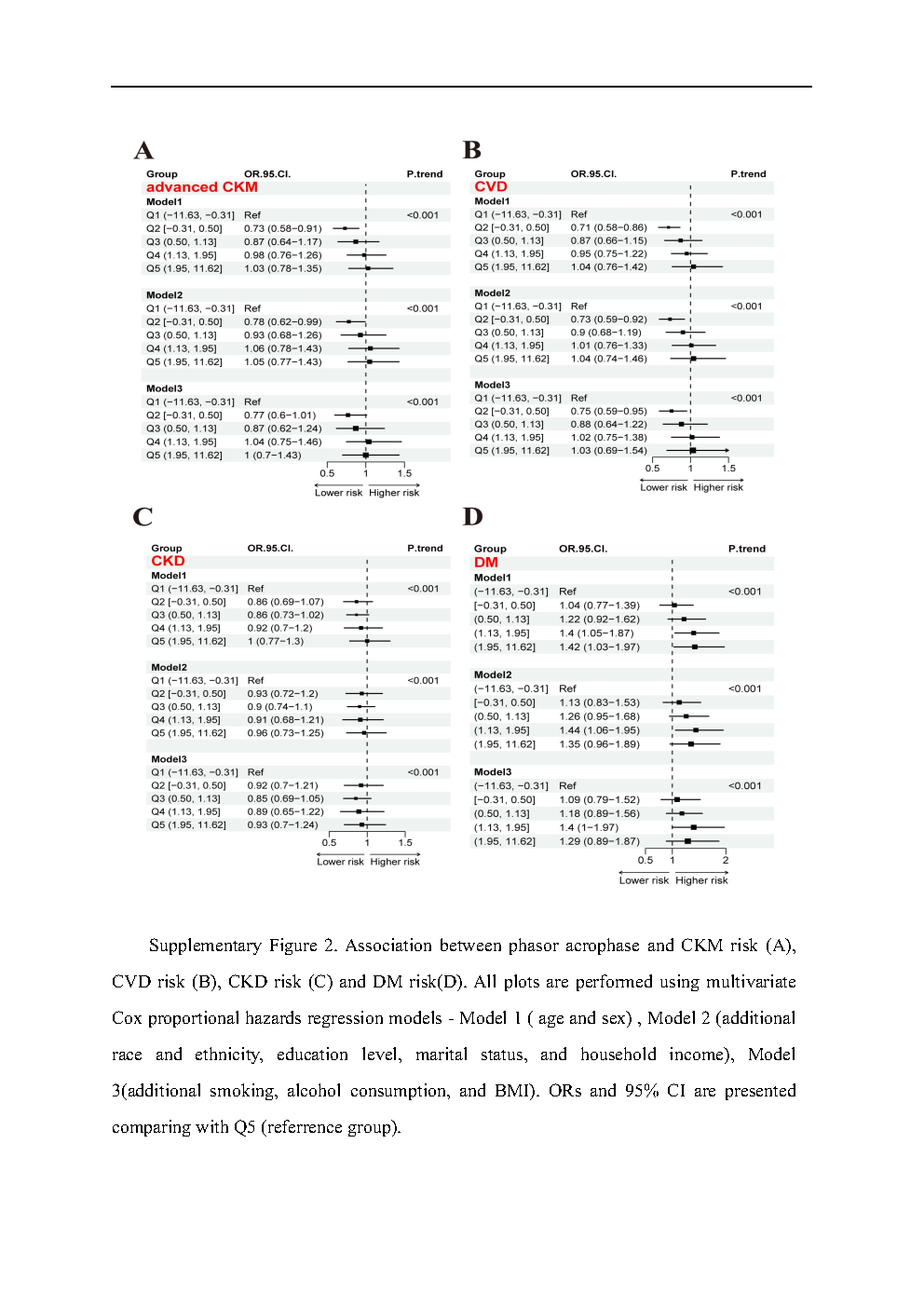

### supplement_Page3.tif

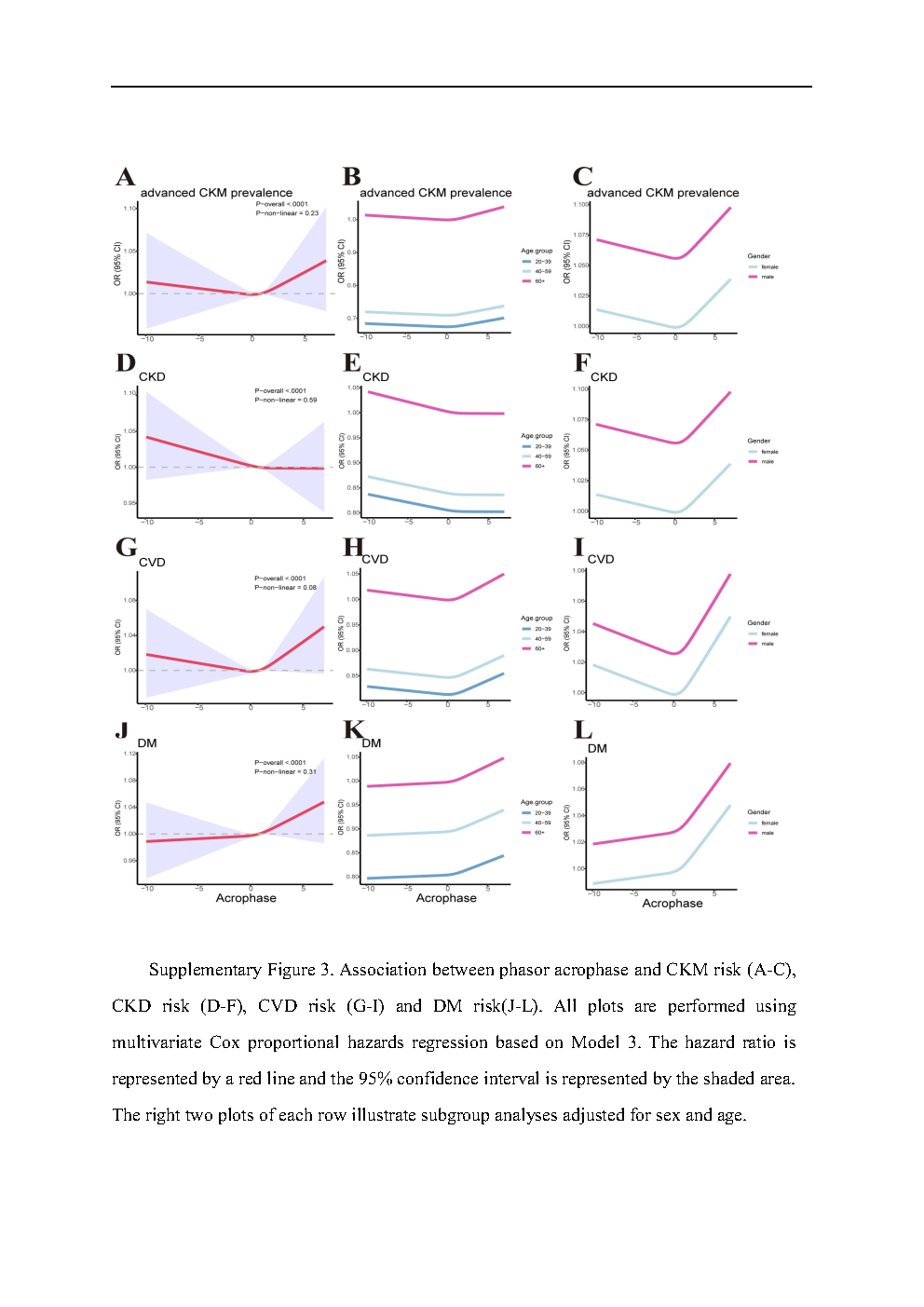

### supplement_Page4.tif

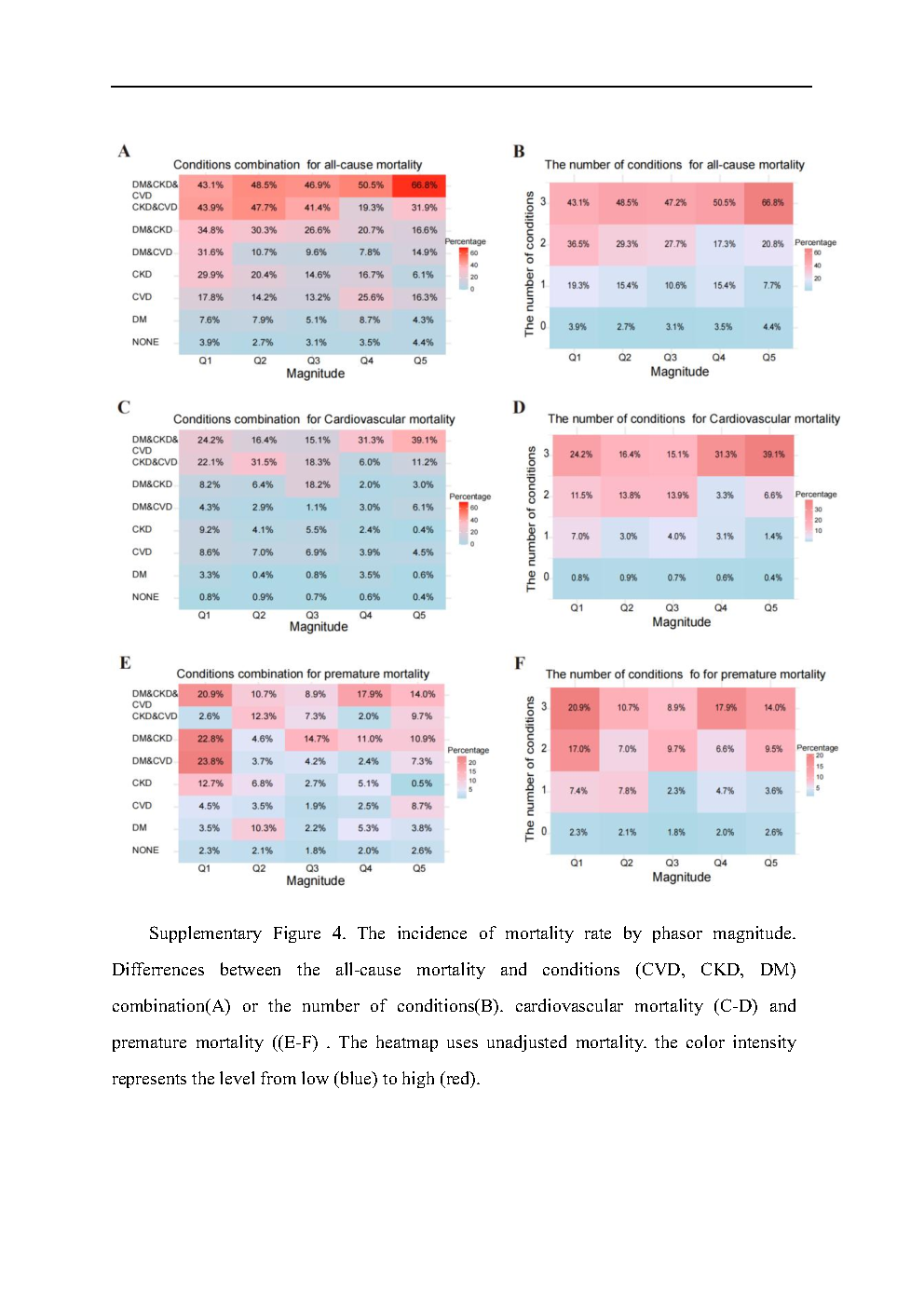

### supplement_Page5.tif

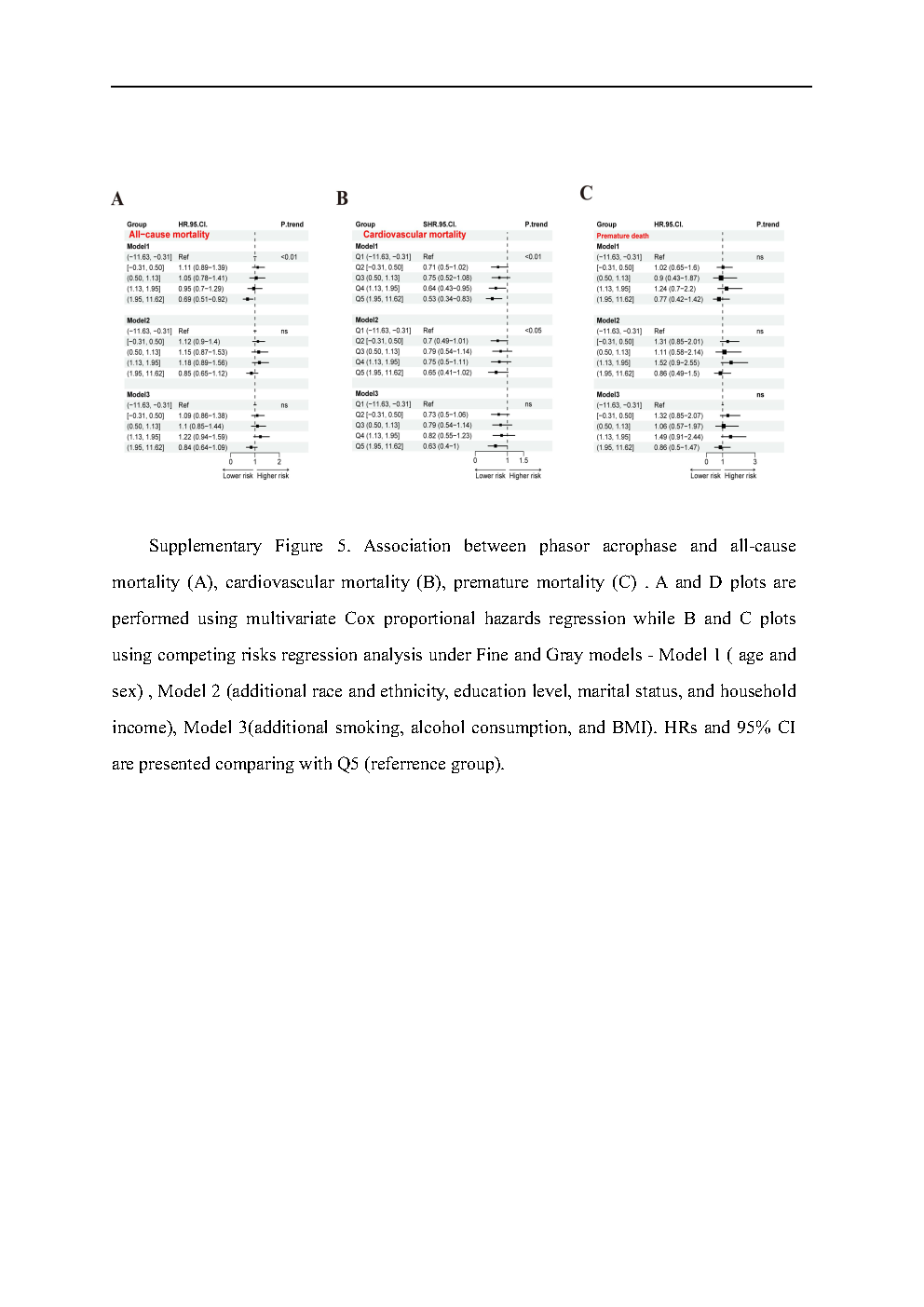

### supplement_Page6.tif

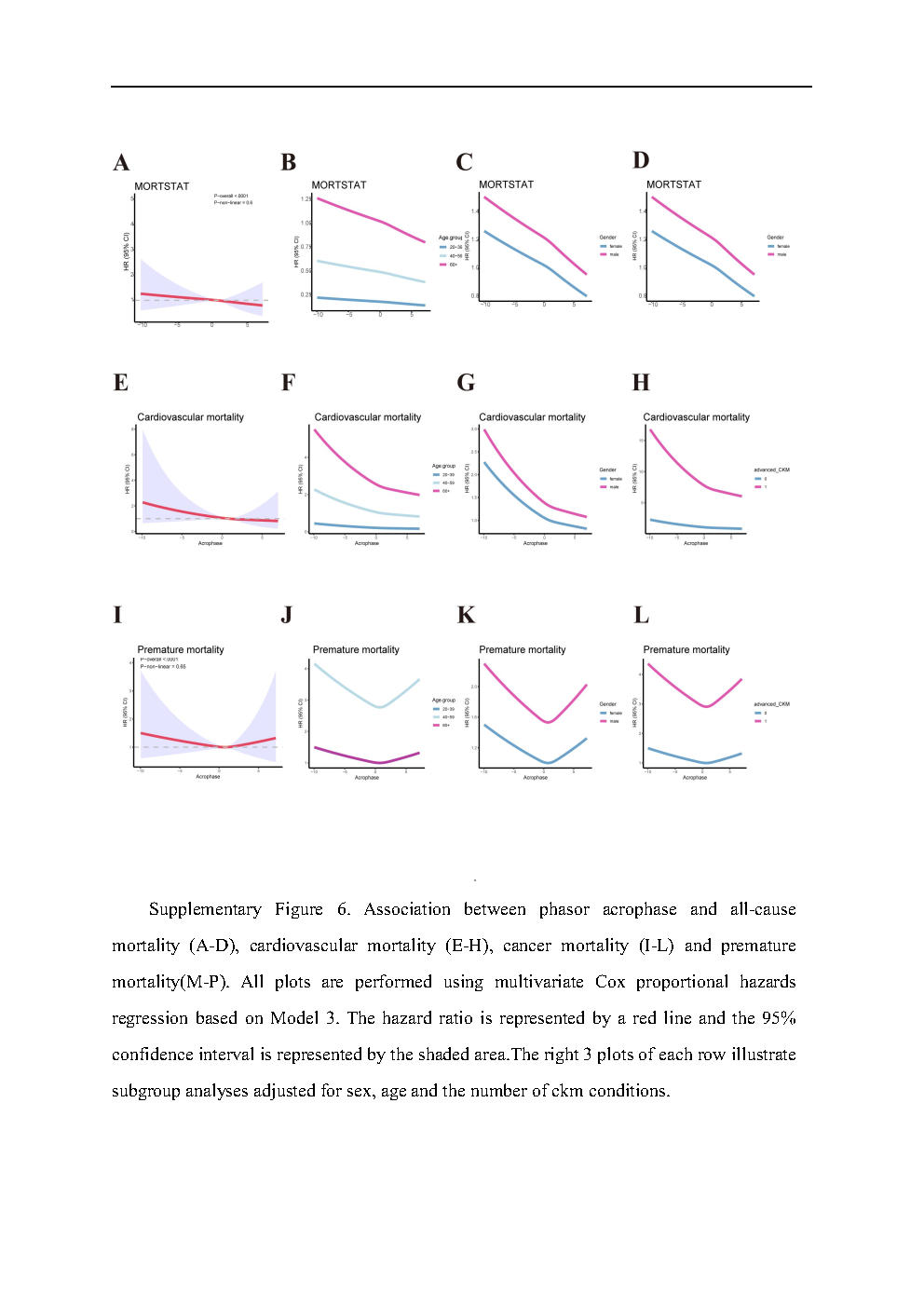
